# Prenatal alcohol consumption and child IQ and cognition: a Mendelian randomization study

**DOI:** 10.1101/2021.10.15.21264993

**Authors:** JA Labrecque, CAM Cecil, SA Swanson

**Author notes:** Corresponsing author.

## Abstract

**Background:** Previous studies using Mendelian randomization have found that fetal alcohol exposure may be associated with lower IQ and test scores in childhood.

**Objectives:** We aim to replicate and extend these findings in Generation R Study, a birth cohort based in Rotterdam, the Netherlands.

**Methods:** We used data from Generation R which recruited pregnant women between 2002 and 2006. Alcohol use was assessed via questionnaire during each trimester. IQ was measured in the children between ages 5 and 8 using the Snijders-Oomen Non-Verbal Intelligence Test. Scores from a national standardized test administered around age 12 were used as a measure of cognition. We estimated the associations between ten genetic variants in the mothers previously found to be related to alcohol consumption and metabolism and each of the outcomes. In the children, we also estimated the association between the same genetic variants as well as two polygenic scores for alcohol consumption and the outcomes.

**Results:** Maternal genetic variants were not found to be related to either outcome but wide confidence intervals did not preclude important effects. A few genetic variants in the children were suggestive of a decrease in IQ. Likewise, one genetic variant and the genetic score had estimates and confidence intervals consistent with increases in standardized test scores.

**Conclusions:** Our results provide slight support for associations between genetic variants in children related to maternal prenatal alcohol consumption and IQ and cognition outcomes. These findings are in line with two previous studies on this topic.

## Introduction

While heavy drinking during pregnancy adversely affects offspring health, less is known about the effects of light to moderate drinking during pregnancy. Studies relying on confounder adjustment have had varied conclusions, ranging from possible adverse^1,2^ to positive^3,4^ effects of light to moderate drinking on cognitive outcomes but unmeasured confounding remains a concern. Two studies using the The Avon Longitudinal Study of Parents and Children (ALSPAC) birth cohort, Lewis et al 2012^5^ and Zuccolo et al 2013^6^ used Mendelian randomization to examine this question. They concluded that alcohol consumption in the first trimester was related to lower IQ at age 8^5^ and worse performance on standardized tests at age 11.^6^ Mendelian randomization can be a good complementary source of evidence but, to date, these findings have not been replicated in an independent cohort. Furthermore, some of the main conclusions of these studies stratify on alcohol consumption, the exposure of interest, which, in Mendelian randomization analyses, can lead to collider bias^7^ potentially inducing a non-causal relationship between genetic variants related to alcohol consumption and child outcomes in the stratified analyses. Here, we replicate and extend the findings of Lewis et al 2012 and Zuccolo et al 2013 using a highly comparable birth cohort from the Netherlands.

## Methods

Generation R is a population-based birth cohort in Rotterdam, the Netherlands. Pregnant women were recruited between April 2002 and January 2006 (N=9778). We restricted to children with two European parents (self-report) to reduce population stratification (N=4285). More information can be found on Generation R elsewhere.^8^

Questionnaires were sent to the participants during early pregnancy including questions about alcohol consumption. Mothers were asked about their alcohol consumption within the past 2 or 3 months and pre-pregnancy.^9^ On average, an alcoholic drink contains 12g of alcohol in the Netherlands.

Two components of a Dutch nonverbal intelligence test, the Snijders-Oomen Niet-Verbale Intelligentietest, were used to assess child IQ between 5 to 8 years of age. The results from the two components were age standardized and converted to non-verbal IQ scores with a mean of 100 and a standard deviation of 15.

In the Netherlands, standardized tests developed by the Central Institute for Test Development (CITO; www.cito.com) are administered around age 12. The CITO test scores range between 501 and 550 points and are used to recommend which type of secondary school is best for each student. The test is not compulsory but 85% of Dutch primary schools participate in the exams. CITO scores were standardized within each year and the resulting distribution was given the same standard distribution as IQ.

We selected the 10 SNPs used in Lewis et al 2012 (which included rs1229984, the SNP used in Zuccolo et al. 2013). More detail on genotyping and quality control for Generation R can be found elsewhere.^10^ In brief, at the time of the current study, genome-wide data on the mothers was available in a subset of 1530 including only 592 European mothers with a European partner. Two SNPs (rs284779 and rs2866151) were available in a larger subgroup. In the children, genetic information was available in 5,732 children.

For each of the 10 genetic variants, we estimated their association with alcohol consumption pre-pregnancy and in the first trimester as was done in Zuccolo et al 2009,^11^ the study which informed the selection of genetic variants in Lewis et al and Zuccolo et al. Briefly, for pre-pregnancy alcohol consumption we regressed categories of alcohol consumption on each genetic variants with ordinal regression. For first trimester alcohol consumption, we regressed alcohol consumption dichotomized as one or more drink per week using logistic regression. We carried out these regressions with all mothers included in our main analyses but also excluding those who did not drink in order to compare with Zuccolo et al 2009.^11^

If, for each genetic variant, the instrumental variables assumptions are satisfied, then the genetic variant’s association with the outcome can be used to test the sharp causal null hypothesis of the effect of alcohol consumption on the outcome. To satisfy these assumptions, the genetic variants must 1) be related to alcohol consumption during the first trimester, 2) not have any effect on the outcome other than through its effect on alcohol consumption during pregnancy and 3) be independent of the counterfactual outcome (i.e. not confounded with the outcome or related through selection bias). As in Lewis et al, we used linear regression to estimate the per allele association between each genetic variant individually and each outcome, IQ and CITO score. Heterozygotes and homozygotes were grouped together for rs1229984 as a previous study suggested a dominant effect.^11^ As a sensitivity analysis, we also used tobit regression with the CITO scores because of a ceiling effect in the CITO data. We ran these analyses for all genetic variants measured in mothers and in children. In another sensitivity analysis, we adjusted the regressions in the children for the same genetic variant measured in the mothers.

We also reconstructed the genetic score used in Lewis et al by adding the number of rare alleles in rs284779, rs4147536, rs975833 and rs2866151. In an additional analysis, we created a genetic score using all 10 variants. We then regressed both outcomes on each of these genetic scores.

In prior ALSPAC publications, many of the analyses of interest were carried out stratified on the alcohol consumption and/or excluding mothers who drank >6 units per week at any point in the pregnancy. This is known to cause collider bias^7^ (Supplementary Material: Figure S1). Therefore, we chose to not replicate these analyses focusing on unstratified results instead.

**Table 1:** Distribution of potential confounders of the relationship between prenatal alcohol consumption and IQ or CITO scores across categories of alcohol consumption. Values are either mean (standard deviation) or count (percentage).

|  | No drinking (N=801) | <1 drink per week (N=680) | 1+ drink per week (N=623) |
| --- | --- | --- | --- |
| Mother’s age in years | 30.9 (4.2) | 31.9 (3.8) | 32.6 (3.8) |
| Finished secondary education | 636 (80.0%) | 625 (92.6%) | 587 (94.5%) |
| Married | 413 (51.6%) | 329 (48.6%) | 279 (44.9%) |
| Primiparous | 479 (59.9%) | 415 (61.0%) | 397 (63.7%) |
| Smoked during 1st trimester | 122 (15.4%) | 149 (22.2%) | 186 (30.0%) |
| Depressed | 24 (3.4%) | 20 (3.3%) | 23 (4.0%) |
| Calcium (mg/day) | 1155.3 (439.6) | 1226.3 (410.2) | 1269.7 (419.5) |
| Vitamin C (mg/day) | 125.7 (60.4) | 130.0 (56.4) | 133.0 (55.6) |
| Iron (mg/day) | 11.5 (3.4) | 12.2 (3.3) | 12.5 (3.3) |
| Folate (nmol/L) | 20.5 (9.4) | 19.8 (8.2) | 21.2 (8.5) |

## Results

Sample sizes were mostly limited by the number of genotyped mothers and children. There was also missingness in IQ and CITO scores due to loss to follow-up ranging between 23-64% (Supplementary Material: Table S1).

Drinking behavior during the first trimester was related to many potential confounders of the relationship between prenatal alcohol consumption and child IQ and test scores. Mothers who consumed more alcohol in the first trimester were older, more likely to be primiparous and more likely to have smoked during the first trimester among other differences. There were no important differences in these variables when comparing mothers with different numbers of alleles of the genetic variants selected (Supplementary Material: Table S2-S11).

The mothers’ genetic variants were at most weakly associated with alcohol consumption during the first trimester (Table 2). In the two genetic variants in the mothers for which we had a larger sample size, we found similar associations with alcohol consumption during the first trimester as was found in ALSPAC.^11^ Among the remaining genetic variants, some point estimates differed from those found in ALSPAC but with wide confidence intervals indicating that this may be due to chance.

**Table 2:** Odds ratio (OR) and 95% confidence interval of the relationship between each SNP and whether the mothers had one or more drink per week in the Generation R cohort and ALSPAC from Zuccolo 2009. Pre-pregnancy odds ratios are from an ordinal regression where alcohol is classified into three categories: <1 drink per week, 1-6 drinks per week and 7+ drinks per week. First trimester odds ratios are from a logistic regression where alcohol consumption was dichotomized as <1 drink per week and 1 or more drink per week.

| SNP | Generation R |  | ALSPAC |
| --- | --- | --- | --- |
|  | All mothers | Only drinkers | Only drinkers |
| **Pre-pregnancy** |  |  |  |
| rs4699714 | 1.09 (0.86, 1.39) | 0.89 (0.68, 1.16) | 1.07 (0.99, 1.15) |
| rs3762894 | 1.17 (0.86, 1.58) | 1.11 (0.79, 1.56) | 0.96 (0.88, 1.05) |
| rs4148884 | 0.79 (0.54, 1.17) | 0.92 (0.59, 1.42) | 1.00 (0.89, 1.13) |
| rs2866151 | 1.04 (0.95, 1.14) | 1.08 (0.97, 1.20) | 1.10 (1.03, 1.17) |
| rs975833 | 1.12 (0.87, 1.45) | 1.06 (0.79, 1.41) | 0.97 (0.90, 1.04) |
| rs1229966 | 1.16 (0.93, 1.46) | 1.11 (0.86, 1.43) | 0.93 (0.87, 0.99) |
| rs2066701 | 1.12 (0.88, 1.43) | 1.08 (0.83, 1.42) | 0.95 (0.88, 1.02) |
| rs4147536 | 0.85 (0.64, 1.13) | 0.95 (0.69, 1.30) | 0.93 (0.86, 1.01) |
| rs1229984 | 0.73 (0.35, 1.50) | 1.26 (0.57, 2.78) | 0.69 (0.56, 0.86) |
| rs284779 | 0.95 (0.86, 1.04) | 0.97 (0.87, 1.07) | 1.03 (0.96, 1.10) |
| **First trimester** |  |  |  |
| rs4699714 | 1.09 (0.82, 1.46) | 1.01 (0.95, 1.08) | 1.01 (0.91, 1.12) |
| rs3762894 | 1.00 (0.69, 1.44) | 0.98 (0.90, 1.06) | 1.04 (0.91, 1.19) |
| rs4148884 | 1.12 (0.71, 1.76) | 1.04 (0.94, 1.15) | 0.99 (0.83, 1.18) |
| rs2866151 | 0.98 (0.87, 1.09) | 1.00 (0.97, 1.02) | 0.99 (0.90, 1.10) |
| rs975833 | 0.84 (0.61, 1.16) | 0.95 (0.89, 1.01) | 1.07 (0.95, 1.20) |
| rs1229966 | 0.86 (0.65, 1.14) | 0.96 (0.91, 1.02) | 1.02 (0.92, 1.13) |
| rs2066701 | 0.78 (0.57, 1.06) | 0.94 (0.88, 1.00) | 1.01 (0.91, 1.13) |
| rs4147536 | 0.77 (0.54, 1.10) | 0.97 (0.90, 1.05) | 0.98 (0.87, 1.10) |
| rs1229984 | 1.05 (0.45, 2.43) | 1.06 (0.87, 1.28) | 0.78 (0.53, 1.13) |
| rs284779 | 1.05 (0.94, 1.18) | 1.02 (0.99, 1.04) | 1.08 (0.98, 1.20) |

Maternal SNPs were weakly associated with child IQ, albeit with wide confidence intervals (Table 3). In the two SNPs for which we had a larger sample size, the confidence intervals were consistent with an effect smaller than one unit (in either direction). The children’s SNPs were likewise weakly associated with child IQ, though the larger sample sizes due to available genetic data led to narrower confidence intervals. One genetic variant, rs4699714, was slightly more strongly, negatively associated with IQ as was the 10 variant genetic score.

**Table 3:**
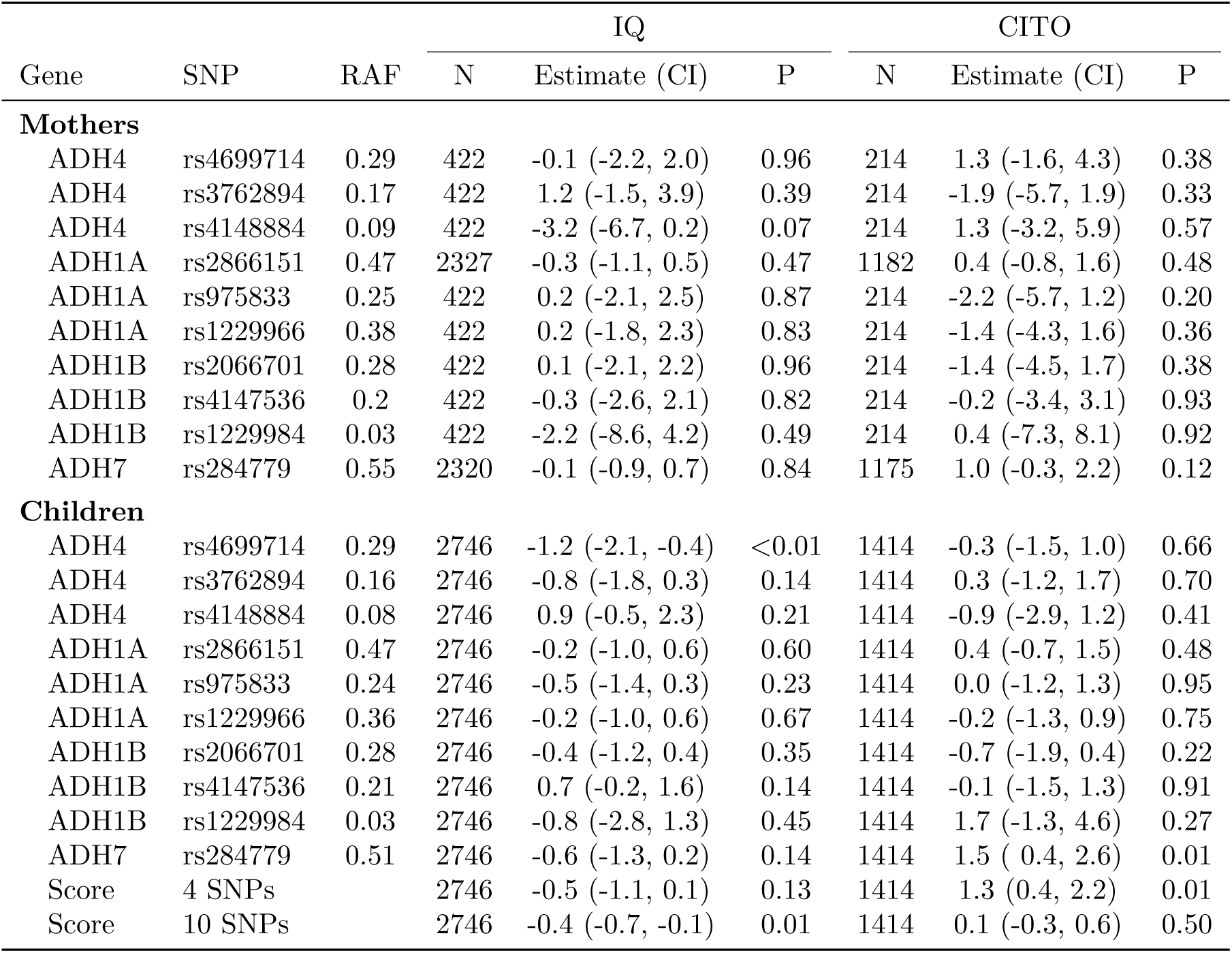
Association between genetic variants measured in mothers and children and IQ as measured by Snijders-Oomen non-verbal intelligence test between age 5 and 8 and CITO scores at age 12. In children, the association between two genetic scores and IQ and CITO scores are also presented.

Maternal SNPs were also weakly associated with CITO scores, again, with wide confidence intervals. For rs284779, we found an estimate consistent mostly with increased CITO scores. The same genetic variant in children was also associated with higher CITO scores. Though the four variant genetic score was associated with important increases in CITO, the 10 variant score was more precise and consistent with only smaller values in both directions.

## Discussion

Overall, most genetic variants did not demonstrate a strong relationship with either IQ or CITO score. There was a slight suggestion of a possible small negative relationship between the genetic variants in children and IQ as was found by Lewis et al 2012. For test scores, our result for the association between maternal rs1229964 and CITO score was consistent with the estimate found by Zuccolo et al 2013 but the confidence intervals were very wide and also consistent with many other possible values meaning that our result does not provide extra evidence for Zuccolo et al 2013’s result. None of the other maternal genetic variants were associated with CITO scores. It should be noted that it is difficult to compare the magnitude of association between the genetic variants and between cohorts because the genetic variants have different relationships with alcohol consumption.

Replication of genetic variant-outcome associations notwithstanding, there are many possible sources of bias that may impact both the current study and the prior ALSPAC results. Though we restricted to children with two European parents, population stratification can bias Mendelian randomization studies. Further, the nature or perinatal epidemiology and prenatal exposures also pose additional sources of selection biases and post-natal “pleiotropy.”^12^ All such biases may be amplified by the observed weak associations between the genetic variants and alcohol consumption during pregnancy. Another limitation of the current study is the relatively small sample sizes for some genetic variants particularly among the mothers.

## Data Availability

All data produced in the present study are available upon reasonable request to the authors.

