## Supplementary Material for "Prenatal alcohol consumption and child IQ and cognition: a Mendelian randomization study"

| **Section** | **Page** |
|---|---|
| Figure S1: Demonstration of collider bias in Mendelian randomization | 2 |
| Table S1: Missingness in IQ and CITO scores | 3 |
| Tables S2-11: Relationship between genetic variants in the mothers and potential confounders of alcohol-outcome relationship | 4-7 |
| Table S12: Sensitivity analysis adjusting for mother's genetic variant | 8 |
| Table S13: Results using tobit regression | 9 |
| Table S14: Unpublished results from Lewis et al 2012 unstratified by alcohol consumption | 10 |

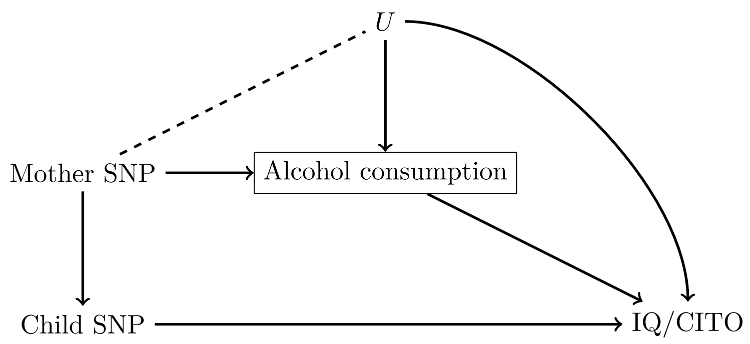


Figure S1: A causal graph depicting the bias potentially induced by stratifying on the mother's alcohol consumption. Stratifying on mother's alcohol consumption (shown as a box) induces a correlation between any SNP that has an effect on alcohol consumption and other variables that are causes of alcohol consumption including confounders of alcohol consumption and the outcomes in this study. Note that stratifying on the mother's alcohol consumption will also open a backdoor path (i.e. a bias path) between the child's SNP and the outcomes considered in this study. Restricting on mother's who did not binge drink will also result in this type of collider bias.

Table S1: Missingness in IQ and CITO scores by genetic variant.

| | | IQ | | | CITO | | |
|---|---|---|---|---|---|---|---|
| Gene | SNP | Missing | Observed | Proportion missing | Missing | Observed | Proportion missing |
| **Mothers** | | | | | | | |
| ADH4 | rs4699714 | 170 | 422 | 0.29 | 378 | 214 | 0.64 |
| ADH4 | rs3762894 | 170 | 422 | 0.29 | 378 | 214 | 0.64 |
| ADH4 | rs4148884 | 170 | 422 | 0.29 | 378 | 214 | 0.64 |
| ADH1A | rs2866151 | 965 | 2327 | 0.29 | 2110 | 1182 | 0.64 |
| ADH1A | rs975833 | 170 | 422 | 0.29 | 378 | 214 | 0.64 |
| ADH1A | rs1229966 | 170 | 422 | 0.29 | 378 | 214 | 0.64 |
| ADH1B | rs2066701 | 170 | 422 | 0.29 | 378 | 214 | 0.64 |
| ADH1B | rs4147536 | 170 | 422 | 0.29 | 378 | 214 | 0.64 |
| ADH1B | rs1229984 | 170 | 422 | 0.29 | 378 | 214 | 0.64 |
| ADH7 | rs284779 | 966 | 2320 | 0.29 | 2111 | 1175 | 0.64 |
| **Children** | | | | | | | |
| ADH4 | rs4699714 | 823 | 2746 | 0.23 | 2155 | 1414 | 0.60 |
| ADH4 | rs3762894 | 823 | 2746 | 0.23 | 2155 | 1414 | 0.60 |
| ADH4 | rs4148884 | 823 | 2746 | 0.23 | 2155 | 1414 | 0.60 |
| ADH1A | rs2866151 | 823 | 2746 | 0.23 | 2155 | 1414 | 0.60 |
| ADH1A | rs975833 | 823 | 2746 | 0.23 | 2155 | 1414 | 0.60 |
| ADH1A | rs1229966 | 823 | 2746 | 0.23 | 2155 | 1414 | 0.60 |
| ADH1B | rs2066701 | 823 | 2746 | 0.23 | 2155 | 1414 | 0.60 |
| ADH1B | rs4147536 | 823 | 2746 | 0.23 | 2155 | 1414 | 0.60 |
| ADH1B | rs1229984 | 823 | 2746 | 0.23 | 2155 | 1414 | 0.60 |
| ADH7 | rs284779 | 823 | 2746 | 0.23 | 2155 | 1414 | 0.60 |

Table S2: Distribution of potential confounders between common heterozygotes and mothers with one rare allele of rs284779.

| | Common homozygote | 1 or 2 Rare allele |
|---|---|---|
| N | 432 | 1659 |
| Age (years) | 31.6 (4.1) | 31.7 (4.0) |
| Finished secondary education | 377 (87.7%) | 1459 (88.5%) |
| Married | 207 (47.9%) | 808 (48.9%) |
| Primiparous | 258 (59.7%) | 1025 (61.8%) |
| Smoked during 1st trimester | 93 (21.5%) | 361 (22.0%) |
| Depressed | 11 (2.9%) | 56 (3.7%) |
| Calcium (mg/day) | 1193.0 (428.9) | 1214.6 (424.8) |
| Vitamin C (mg/day) | 127.8 (59.5) | 129.4 (56.9) |
| Iron (mg/day) | 12.1 (3.5) | 12.0 (3.3) |
| Folate (nmol/L) | 21.0 (9.4) | 20.3 (8.6) |

Table S3: Distribution of potential confounders between common heterozygotes and mothers with one rare allele of rs2866151.

| | Common homozygote | 1 or 2 Rare allele |
|---|---|---|
| N | 589 | 1515 |
| Age (years) | 31.8 (3.9) | 31.7 (4.1) |
| Finished secondary education | 529 (90.1%) | 1319 (87.7%) |
| Married | 292 (49.6%) | 729 (48.3%) |
| Primiparous | 353 (60.0%) | 938 (61.9%) |
| Smoked during 1st trimester | 133 (22.7%) | 324 (21.6%) |
| Depressed | 19 (3.6%) | 48 (3.5%) |
| Calcium (mg/day) | 1214.4 (398.7) | 1212.2 (437.5) |
| Vitamin C (mg/day) | 128.8 (57.1) | 129.5 (58.0) |
| Iron (mg/day) | 12.3 (3.5) | 11.9 (3.3) |
| Folate (nmol/L) | 19.8 (9.0) | 20.8 (8.7) |

Table S4: Distribution of potential confounders between common heterozygotes and mothers with one rare allele of rs4699714.

| | Common homozygote | 1 or 2 Rare allele |
|---|---|---|
| N | 159 | 143 |
| Age (years) | 31.7 (4.3) | 31.6 (4.1) |
| Finished secondary education | 128 (80.5%) | 128 (89.5%) |
| Married | 65 (41.1%) | 66 (46.2%) |
| Primiparous | 93 (58.5%) | 98 (68.5%) |
| Smoked during 1st trimester | 34 (21.7%) | 29 (20.3%) |
| Depressed | 9 (6.4%) | 4 (3.0%) |
| Calcium (mg/day) | 1184.9 (449.1) | 1229.9 (425.7) |
| Vitamin C (mg/day) | 131.7 (60.7) | 130.0 (49.8) |
| Iron (mg/day) | 11.7 (3.1) | 12.6 (3.6) |
| Folate (nmol/L) | 19.4 (8.6) | 21.1 (8.8) |

Table S5: Distribution of potential confounders between common heterozygotes and mothers with one rare allele of rs3762894.

| | Common homozygote | 1 or 2 Rare allele |
|---|---|---|
| N | 207 | 95 |
| Age (years) | 31.4 (4.2) | 32.1 (4.3) |
| Finished secondary education | 176 (85.0%) | 80 (84.2%) |
| Married | 93 (44.9%) | 38 (40.4%) |
| Primiparous | 135 (65.2%) | 56 (58.9%) |
| Smoked during 1st trimester | 39 (19.0%) | 24 (25.3%) |
| Depressed | 10 (5.3%) | 3 (3.6%) |
| Calcium (mg/day) | 1195.4 (431.0) | 1229.8 (453.9) |
| Vitamin C (mg/day) | 131.5 (55.6) | 129.5 (56.1) |
| Iron (mg/day) | 12.1 (3.5) | 12.1 (3.2) |
| Folate (nmol/L) | 20.1 (8.5) | 20.5 (9.2) |

Table S6: Distribution of potential confounders between common heterozygotes and mothers with one rare allele of rs4148884.

| | Common homozygote | 1 or 2 Rare allele |
|---|---|---|
| N | 244 | 58 |
| Age (years) | 31.6 (4.4) | 31.7 (3.6) |
| Finished secondary education | 205 (84.0%) | 51 (87.9%) |
| Married | 100 (41.2%) | 31 (53.4%) |
| Primiparous | 154 (63.1%) | 37 (63.8%) |
| Smoked during 1st trimester | 50 (20.6%) | 13 (22.8%) |
| Depressed | 9 (4.1%) | 4 (7.8%) |
| Calcium (mg/day) | 1236.7 (434.4) | 1083.6 (434.5) |
| Vitamin C (mg/day) | 130.3 (52.2) | 133.4 (68.4) |
| Iron (mg/day) | 12.2 (3.3) | 11.7 (3.6) |
| Folate (nmol/L) | 20.1 (8.8) | 21.0 (8.4) |

Table S7: Distribution of potential confounders between common heterozygotes and mothers with one rare allele of rs975833.

| | Common homozygote | 1 or 2 Rare allele |
|---|---|---|
| N | 176 | 126 |
| Age (years) | 31.5 (4.1) | 31.9 (4.4) |
| Finished secondary education | 151 (85.8%) | 105 (83.3%) |
| Married | 79 (44.9%) | 52 (41.6%) |
| Primiparous | 115 (65.3%) | 76 (60.3%) |
| Smoked during 1st trimester | 37 (21.3%) | 26 (20.6%) |
| Depressed | 8 (5.0%) | 5 (4.4%) |
| Calcium (mg/day) | 1179.5 (417.8) | 1244.1 (463.9) |
| Vitamin C (mg/day) | 130.9 (53.7) | 130.9 (58.7) |
| Iron (mg/day) | 12.0 (3.5) | 12.3 (3.3) |
| Folate (nmol/L) | 19.7 (8.5) | 20.9 (9.0) |

Table S8: Distribution of potential confounders between common heterozygotes and mothers with one rare allele of rs1229966.

| | Common homozygote | 1 or 2 Rare allele |
|---|---|---|
| N | 129 | 173 |
| Age (years) | 31.3 (3.8) | 31.9 (4.5) |
| Finished secondary education | 108 (83.7%) | 148 (85.5%) |
| Married | 55 (42.6%) | 76 (44.2%) |
| Primiparous | 82 (63.6%) | 109 (63.0%) |
| Smoked during 1st trimester | 26 (20.5%) | 37 (21.4%) |
| Depressed | 7 (6.1%) | 6 (3.8%) |
| Calcium (mg/day) | 1164.0 (431.7) | 1238.3 (441.3) |
| Vitamin C (mg/day) | 131.3 (53.7) | 130.6 (57.3) |
| Iron (mg/day) | 11.8 (3.6) | 12.4 (3.2) |
| Folate (nmol/L) | 19.1 (8.6) | 21.0 (8.8) |

Table S9: Distribution of potential confounders between common heterozygotes and mothers with one rare allele of rs2066701.

| | Common homozygote | 1 or 2 Rare allele |
|---|---|---|
| N | 168 | 134 |
| Age (years) | 31.5 (4.0) | 31.8 (4.5) |
| Finished secondary education | 143 (85.1%) | 113 (84.3%) |
| Married | 72 (42.9%) | 59 (44.4%) |
| Primiparous | 109 (64.9%) | 82 (61.2%) |
| Smoked during 1st trimester | 36 (21.7%) | 27 (20.1%) |
| Depressed | 8 (5.3%) | 5 (4.2%) |
| Calcium (mg/day) | 1177.6 (419.6) | 1242.0 (458.8) |
| Vitamin C (mg/day) | 132.6 (57.6) | 128.8 (53.4) |
| Iron (mg/day) | 11.9 (3.5) | 12.4 (3.2) |
| Folate (nmol/L) | 19.7 (8.7) | 20.9 (8.8) |

Table S10: Distribution of potential confounders between common heterozygotes and mothers with one rare allele of rs4147536.

| | Common homozygote | 1 or 2 Rare allele |
|---|---|---|
| N | 197 | 105 |
| Age (years) | 31.9 (4.2) | 31.2 (4.3) |
| Finished secondary education | 170 (86.3%) | 86 (81.9%) |
| Married | 89 (45.2%) | 42 (40.4%) |
| Primiparous | 123 (62.4%) | 68 (64.8%) |
| Smoked during 1st trimester | 40 (20.4%) | 23 (22.1%) |
| Depressed | 8 (4.5%) | 5 (5.3%) |
| Calcium (mg/day) | 1222.2 (443.1) | 1178.5 (429.4) |
| Vitamin C (mg/day) | 130.0 (52.6) | 132.5 (61.0) |
| Iron (mg/day) | 12.2 (3.5) | 12.0 (3.3) |
| Folate (nmol/L) | 20.7 (9.0) | 19.4 (8.1) |

Table S11: Distribution of potential confounders between common heterozygotes and mothers with one rare allele of rs1229984.

| | Common homozygote | 1 or 2 Rare allele |
|---|---|---|
| N | 287 | 15 |
| Age (years) | 31.6 (4.2) | 31.5 (4.0) |
| Finished secondary education | 246 (85.7%) | 10 (66.7%) |
| Married | 127 (44.4%) | 4 (26.7%) |
| Primiparous | 181 (63.1%) | 10 (66.7%) |
| Smoked during 1st trimester | 59 (20.6%) | 4 (28.6%) |
| Depressed | 13 (5.0%) | 0 (0.0%) |
| Calcium (mg/day) | 1208.5 (436.1) | 1166.3 (484.4) |
| Vitamin C (mg/day) | 129.6 (51.7) | 154.0 (105.0) |
| Iron (mg/day) | 12.2 (3.4) | 11.1 (4.0) |
| Folate (nmol/L) | 20.3 (8.5) | 19.6 (12.8) |

Table S12: Sensitivity analysis adjusting the relationship between genetic variants in children for the same genetic variant in the mother.

| | | IQ | | | | CITO | | |
|---|---|---|---|---|---|---|---|---|
| Gene | SNP | RAF | N | Estimate (CI) | p | N | Estimate (CI) | p |
| ADH4 | rs4699714 | 0.29 | 391 | -2.7 (-5.0, -0.5) | 0.02 | 200 | -1.1 (-4.3, 2.2) | 0.52 |
| ADH4 | rs3762894 | 0.16 | 391 | 0.6 (-2.1, 3.3) | 0.66 | 200 | 2.2 (-1.5, 5.9) | 0.24 |
| ADH4 | rs4148884 | 0.08 | 391 | 3.2 (-0.4, 6.8) | 0.08 | 200 | -2.5 (-7.3, 2.3) | 0.31 |
| ADH1A | rs2866151 | 0.47 | 2139 | 0.2 (-0.8, 1.2) | 0.71 | 1096 | 0.2 (-1.3, 1.7) | 0.80 |
| ADH1A | rs975833 | 0.24 | 391 | -1.4 (-3.8, 0.9) | 0.24 | 200 | 2.2 (-1.2, 5.5) | 0.20 |
| ADH1A | rs1229966 | 0.36 | 391 | -0.1 (-2.2, 2.0) | 0.94 | 200 | 1.3 (-1.5, 4.1) | 0.38 |
| ADH1B | rs2066701 | 0.29 | 391 | -0.6 (-2.7, 1.5) | 0.59 | 200 | -0.4 (-3.4, 2.6) | 0.79 |
| ADH1B | rs4147536 | 0.20 | 391 | -0.2 (-2.9, 2.4) | 0.88 | 200 | 0.0 (-4.2, 4.3) | 0.98 |
| ADH1B | rs1229984 | 0.04 | 391 | 2.5 (-2.8, 7.9) | 0.35 | 200 | 0.4 (-6.3, 7.0) | 0.92 |
| ADH7 | rs284779 | 0.52 | 2132 | -1.2 (-2.1, -0.2) | 0.01 | 1091 | 0.8 (-0.6, 2.2) | 0.25 |

Table S13: Associations between genetic variants in the children and CITO in the child at age 12 using tobit regression.

| SNP | RAF | N | Estimate (CI) | p |
|---|---|---|---|---|
| **Mothers** | | | | |
| rs4699714 | 0.29 | 214 | 1.6 (-1.4, 4.7) | 0.29 |
| rs3762894 | 0.17 | 214 | -2.1 (-6.1, 1.8) | 0.29 |
| rs4148884 | 0.09 | 214 | 1.0 (-3.7, 5.7) | 0.67 |
| rs2866151 | 0.47 | 1182 | 0.5 (-0.8, 1.7) | 0.46 |
| rs975833 | 0.25 | 214 | -2.6 (-6.1, 1.0) | 0.16 |
| rs1229966 | 0.38 | 214 | -1.3 (-4.4, 1.7) | 0.39 |
| rs2066701 | 0.28 | 214 | -1.7 (-4.9, 1.6) | 0.31 |
| rs4147536 | 0.2 | 214 | 0.1 (-3.3, 3.5) | 0.97 |
| rs1229984 | 0.03 | 214 | 0.1 (-7.8, 8.0) | 0.98 |
| rs284779 | 0.55 | 1175 | 0.9 (-0.3, 2.2) | 0.15 |
| **Children** | | | | |
| rs4699714 | 0.30 | 1414 | -0.4 (-1.7, 0.8) | 0.50 |
| rs3762894 | 0.16 | 1414 | 0.3 (-1.2, 1.8) | 0.73 |
| rs4148884 | 0.08 | 1414 | -1.0 (-3.0, 1.1) | 0.37 |
| rs2866151 | 0.48 | 1414 | 0.4 (-0.8, 1.5) | 0.54 |
| rs975833 | 0.24 | 1414 | 0.0 (-1.3, 1.2) | 0.94 |
| rs1229966 | 0.36 | 1414 | -0.3 (-1.4, 0.9) | 0.66 |
| rs2066701 | 0.29 | 1414 | -0.8 (-2.0, 0.4) | 0.19 |
| rs4147536 | 0.20 | 1414 | 0.1 (-1.4, 1.5) | 0.94 |
| rs1229984 | 0.04 | 1414 | 1.6 (-1.4, 4.7) | 0.29 |
| rs284779 | 0.54 | 1414 | 1.6 ( 0.4, 2.7) | 0.01 |
| Score (4 SNPs) | | 1414 | 1.3 (0.4, 2.3) | 0.09 |
| Score (10 SNPs) | | 1414 | 0.1 (-0.3, 0.6) | 0.01 |

Table S14: Associations between genetics variants and child IQ as measured by the Weschler Intelligence Scale for Children. These results did not appear in Lewis et al 2012[1] (only results stratified by the mother's alcohol consumption were reported) but the authors were extremely kind providing us with these results and allowing us to publish them in supplementary material.

| Gene | SNP | RaF | N | Estimate (CI) | p |
|---|---|---|---|---|---|
| **Mothers** | | | | | |
| ADH4 | rs4699714 | 0.27 | 3916 | -0.17 (-0.97, 0.63) | 0.68 |
| ADH4 | rs3762894 | 0.17 | 3913 | 0.26 (-0.70, 1.22) | 0.60 |
| ADH4 | rs4148884 | 0.09 | 3926 | -1.19 (-2.46, 0.08) | 0.07 |
| ADH1A | rs2866151 | 0.46 | 3850 | -0.50 (-1.23, 0.23) | 0.18 |
| ADH1A | rs975833 | 0.24 | 3880 | 0.35 (-0.51, 1.21) | 0.43 |
| ADH1A | rs1229966 | 0.37 | 3848 | 0.62 (-0.14, 1.38) | 0.11 |
| ADH1B | rs2066701 | 0.29 | 3827 | 0.21 (-0.59, 1.01) | 0.62 |
| ADH1B | rs4147536 | 0.22 | 3884 | 0.26 (-0.62, 1.14) | 0.56 |
| ADH1B | rs1229984 | 0.03 | 3911 | 0.43 (-1.92, 2.78) | 0.72 |
| ADH7 | rs284779 | 0.45 | 3917 | -0.69 (-1.42, 0.04) | 0.06 |
| **Children** | | | | | |
| ADH4 | rs4699714 | 0.27 | 4962 | -0.74 (-1.47, -0.01) | 0.04 |
| ADH4 | rs3762894 | 0.17 | 4849 | 0.18 (-0.68, 1.04) | 0.68 |
| ADH4 | rs4148884 | 0.08 | 4885 | 0.96 (-0.22, 2.14) | 0.11 |
| ADH1A | rs2866151 | 0.47 | 4783 | -0.14 (-0.77, 0.49) | 0.67 |
| ADH1A | rs975833 | 0.24 | 4826 | -0.31 (-1.07, 0.45) | 0.42 |
| ADH1A | rs1229966 | 0.36 | 4844 | 0.24 (-0.43, 0.91) | 0.48 |
| ADH1B | rs2066701 | 0.29 | 4754 | -0.03 (-0.74, 0.68) | 0.93 |
| ADH1B | rs4147536 | 0.21 | 4852 | -0.13 (-0.93, 0.67) | 0.75 |
| ADH1B | rs1229984 | 0.03 | 5574 | 0.34 (-1.62, 2.30) | 0.73 |
| ADH7 | rs284779 | 0.45 | 4868 | -0.78 (-1.43, -0.13) | 0.02 |

[1] Lewis SJ, Zuccolo L, Davey Smith G, Macleod J, Rodriguez S, Draper ES, et al. Fetal AlcoholExposure and IQ at Age 8: Evidence from a Population-Based Birth-Cohort Study. *PLoS ONE* 2012;7.
